# Prediction of Sex and Age from Macular Optical Coherence Tomography Images and Feature Analysis Using Deep Learning

**DOI:** 10.1101/2020.12.23.20248805

**Authors:** Kuan-Ming Chueh, Yi-Ting Hsieh, Homer H. Chen, I-Hsin Ma, Sheng-Lung Huang

## Abstract

The prevalence of certain macular diseases differs between male and female. However, the actual difference in macular structure between male and female was barely understood. Previous studies reported the mean retinal thickness of macula was thinner for female, but here it was observed that the difference is not statistically large enough for sex distinction. Similarly, the age-related non-pathological change of macular structure was also hardly known. It has been found that the thickness of choroid decreases with age. In this study, deep learning was applied to distinguish sex and age from macular optical coherence tomography (OCT) images of 3134 persons and achieved a sex prediction accuracy of 85.6 ± 2.1% and an age prediction error of 5.78 ± 0.29 years. A thorough analysis of the prediction accuracy and the Grad-CAM showed that 1) the foveal contour leads to a better sex distinction than the macular thickness, 2) B-scan macular OCT images contain more sex-related information than en face fundus images, and 3) the age-related characteristics of the macula are on the whole layers of the retina, not just the choroid. These novel findings reported in this study are useful to ophthalmologists for further investigation in the pathogenesis of sex and age-related macular structural diseases.

## Introduction

The macula is located at the center of the retina. It is the area with the densest photoreceptor cone cells and is responsible for the central vision^1^. In the past, sexual difference in the structure of the macula was not known, except few studies have shown that the average thickness of the macula in men was thicker than in women^2-7^. In 2018, Poplin et al.^8^ reported that their deep learning model could distinguish the sex of the subject from color fundus photography, with 97% accuracy. This means that some differences do exist between male and female at the posterior pole including the macula and optic disc, but such differences cannot be easily identified by ophthalmologists. The fovea is a pitted structure at the center of the macula. The shape of the foveal pit could differ a lot among normal populations, and such variation has no special clinical significance^9^. Recently, a study by our team has found that a wide-based foveal pit, which was demonstrated on optical coherence tomography (OCT), occurred five times as frequently in female as in male. Eyes with a wide-based foveal pit tended to have epiretinal membrane formation, and their contralateral eyes also had an extremely high proportion of epiretinal membrane and macular hole^10^. Interestingly, these macular structural diseases including epiretinal membrane and macular hole, also occur more frequently in women^11,12^. In the past, the ophthalmologists had no special hypothesis about the sexual differences in these macular structural diseases. According to the above-mentioned observations, we speculate some inherent differences in the anatomical structure of the macula between male and female, which result in their differences in the prevalence of structure-related maculopathy. Idiopathic epiretinal membrane and macular hole also have age predilection; with higher prevalence noted in middle-aged and elderly people^13-15^. In addition to vitreous degeneration and cell aging, it is still not clear whether the macula itself has age-related structural changes that make the macula predisposes to these lesions. Therefore, it is clinically important to study the sex and age-related differences in the structure of the macula.

Deep learning allows an algorithm to learn the appropriate features by multiple computation layers rather than requiring features to be hand engineered^16^. Some study groups have used deep learning to do the structural segmentation for macular OCT and classification of macular diseases using macular OCT^17-22^. However, deep learning has never been used to predict sex and age through macular OCT images. Recently, gradient-weighted class activation mapping (Grad-CAM) has been widely adopted which uses the gradient information flowing into the final convolutional layer of a convolutional neural network (CNN) to produce a localization map of the important regions in the image^23^. In this study, deep learning to train CNN was used to predict sex and age according to macular OCT images, and then Grad-CAM was used to explore the sex and age-related features from different layers of CNNs in order to offer a new understanding of the macular structure.

## Results

Deep-learning models were developed by using the 6-mm cross-sectional (B-scan) OCT images of the macula. These images were extracted from the macula volume scan using Heidelberg Spectralis (Heidelberg Engineering, Heidelberg, Germany). Each volume scan contained 25 horizontal B-scans that covered the 6 x 6 mm^2^ area of the central macula. Since the most inferior one could not be obtained for the dataset, 24 macular OCT B-scan images were obtained from each set of volume scan (Figure 1). The sex-prediction models were developed using 4866 sets of OCT images (50% of them were female) from the eyes of 2288 persons, and the age-prediction models were developed using 6147 sets of OCT images (average age: 55.2 ± 15.0 years) from the eyes of 3134 persons. Except for using the total 24 images from the volume scan as the input, the 24 images were also divided into 7 groups to analyze from different position of the macula, which included: (1) the 11^th^ image only, which passed through the center of the fovea, (2) the 10^*th*^ and 12^*th*^ images, which passed through the inferior and superior margins of the 500 μm-diameter circle of the fovea, (3) the 9^*th*^ and 13^*th*^ images, which passed through the inferior and superior margins of the 1000 μm-diameter circle of the fovea, (4) the 0^*th*^ - 8^*th*^ and 14^*th*^-23^*th*^ images, (5) the 0^*th*^ and 22^*th*^ images, (6) the 11^*th*^ and 12^*th*^ images, and (7) the total 24 images.

**Figure 1.**
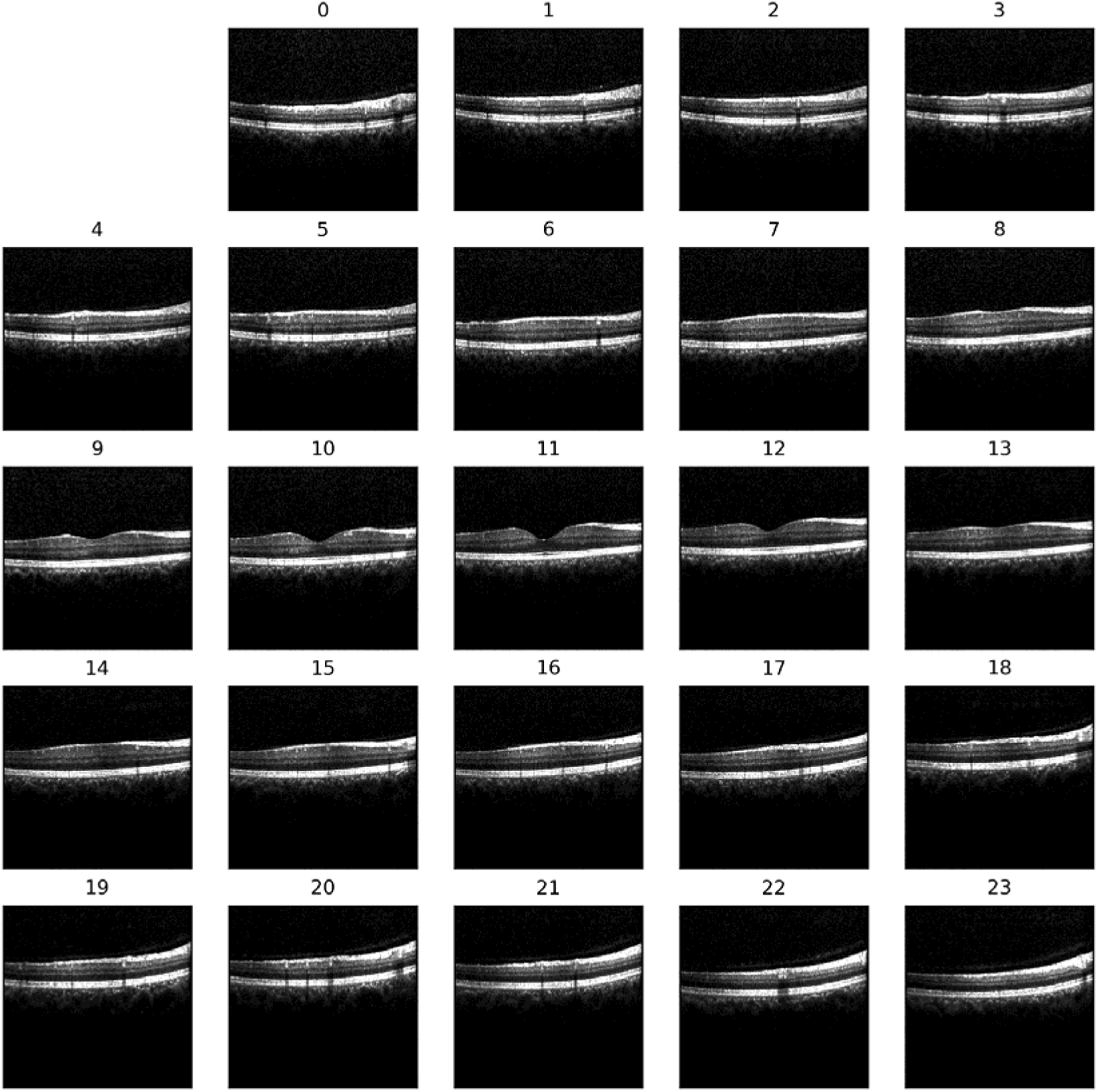
24 images (1 volumes) scanned by Heidelberg Spectralis OCT. The numbers defined in a single volume are shown on the top of each image. Each image contains 496 512 pixels, and the pixel resolution are 3.9 and 11 in the axial and lateral directions, respectively.

### Sex prediction by deep learning models using macular OCT images

The deep-learning models were trained and validated by the 10-fold cross-validation with a variety of input data. The test accuracies are shown in Table 1. The accuracy of sex prediction among different positions of the macula were compared among four models that contained the same amount of input data from different scanning positions, which were (A) 11^*th*^ and 12^*th*^, (B) 10^*th*^ and 12 ^*th*^, (C) 9 ^*th*^ and 13 ^*th*^, (D) 0 ^*th*^ and 22 ^*th*^. These data groups contained OCT images from two different scanning positions that were symmetric to central fovea except for (A), which contained one image in sets passing through the central fovea and the other image in sets passing though the superior juxtafovea. The distance to fovea was the shortest in (A) and the longest in (E). The average accuracy in sex prediction for models(A), (B), (C) and (D) were 75.9 ± 2.3%, 74.8 ± 3.1%, 73.7 ± 2.5% and 70.5 ± 3.0%, respectively. The accuracy decreased gradually when the scanning positions were getting away from the central fovea. When using the 11^*th*^ images only for training and prediction in (E), the accuracy was only 73.1 ± 1.6%, which was less than that in (A). When using all 19 extrafoveal images in (F) or mixed the fovea-containing and extrafoveal images (24 images in sets) together in (G), the accuracy increased to 77.1 ± 2.1% and 77.4 ± 2.2%, respectively.

**Table 1.**
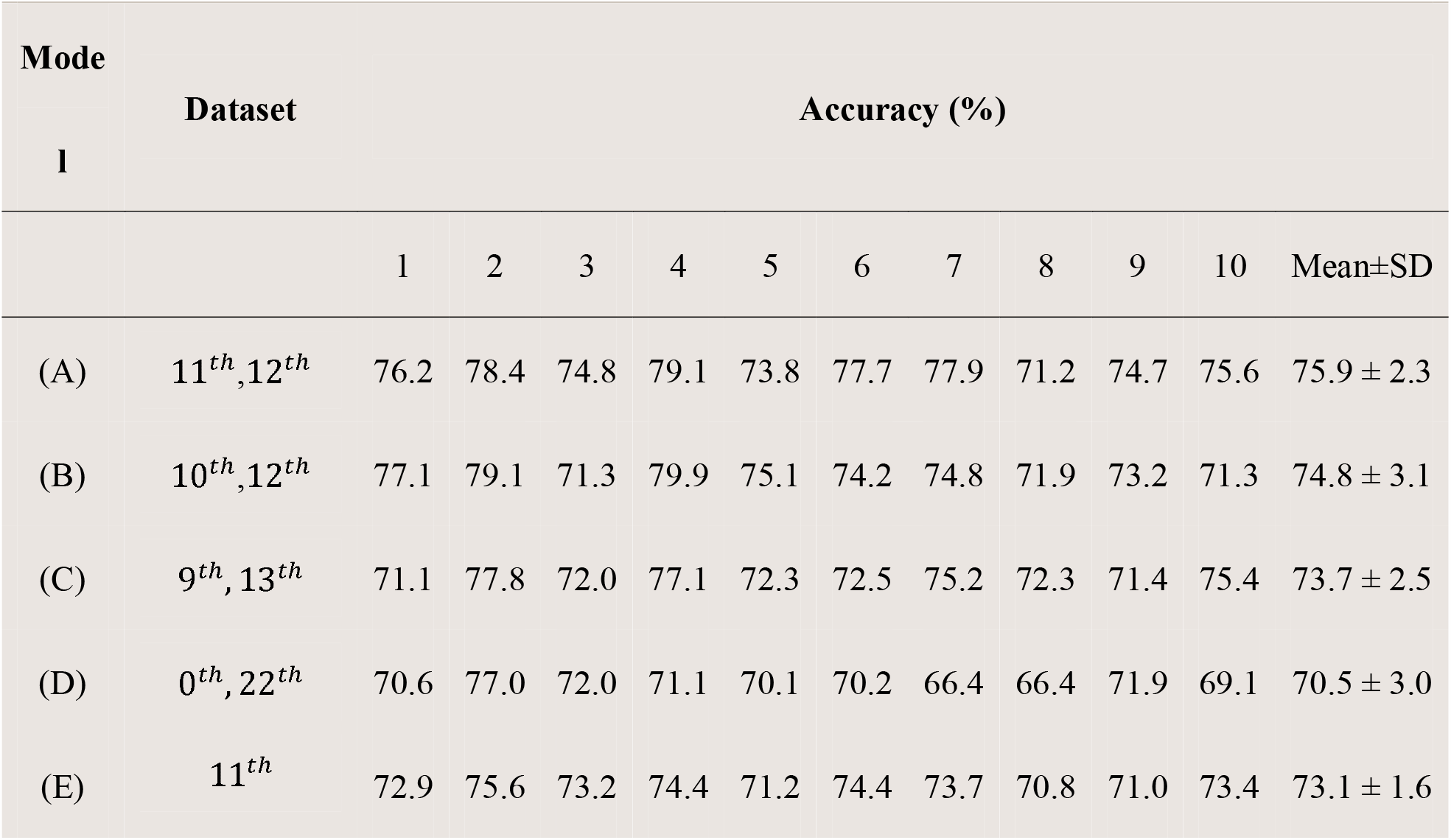
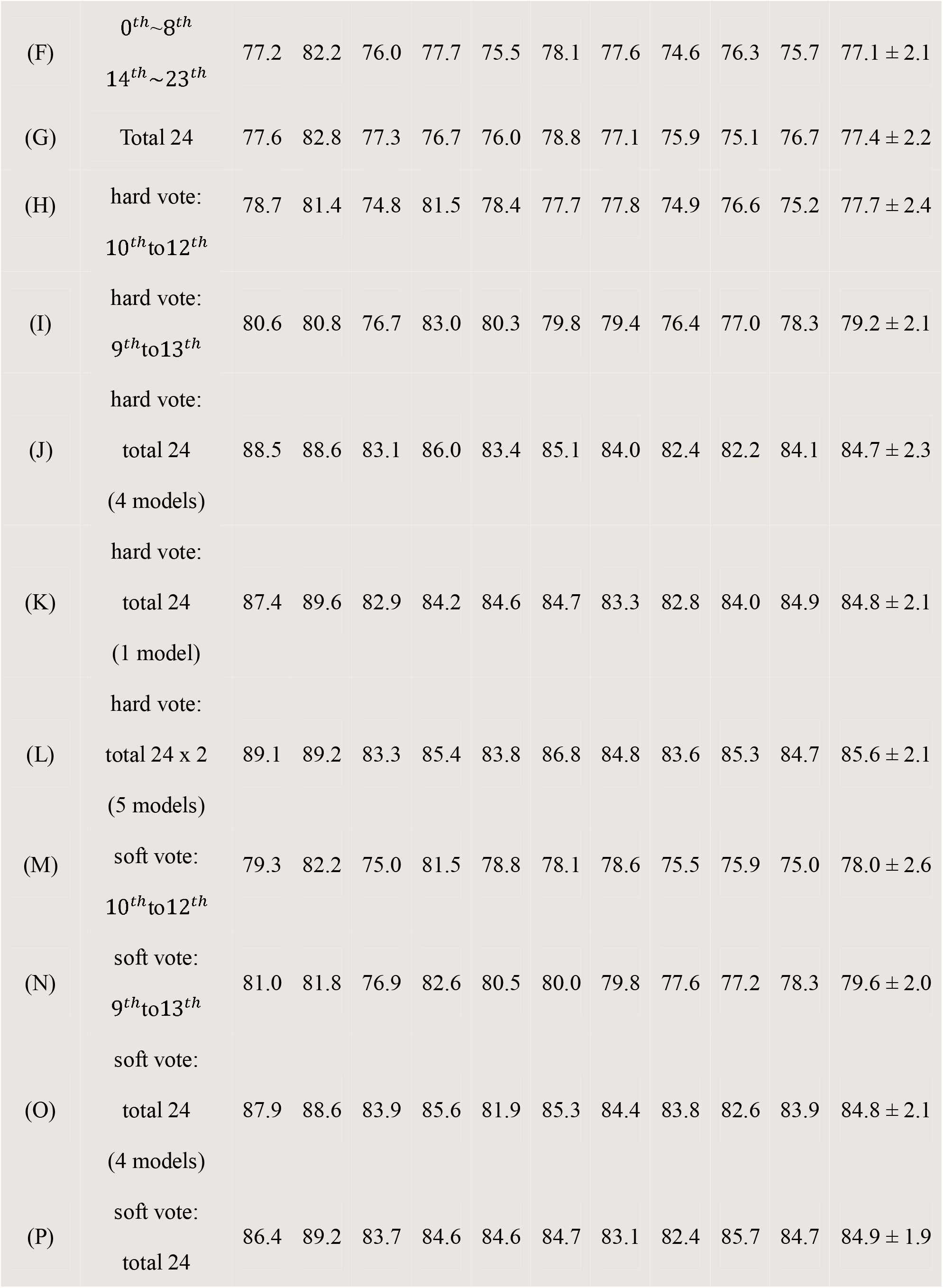
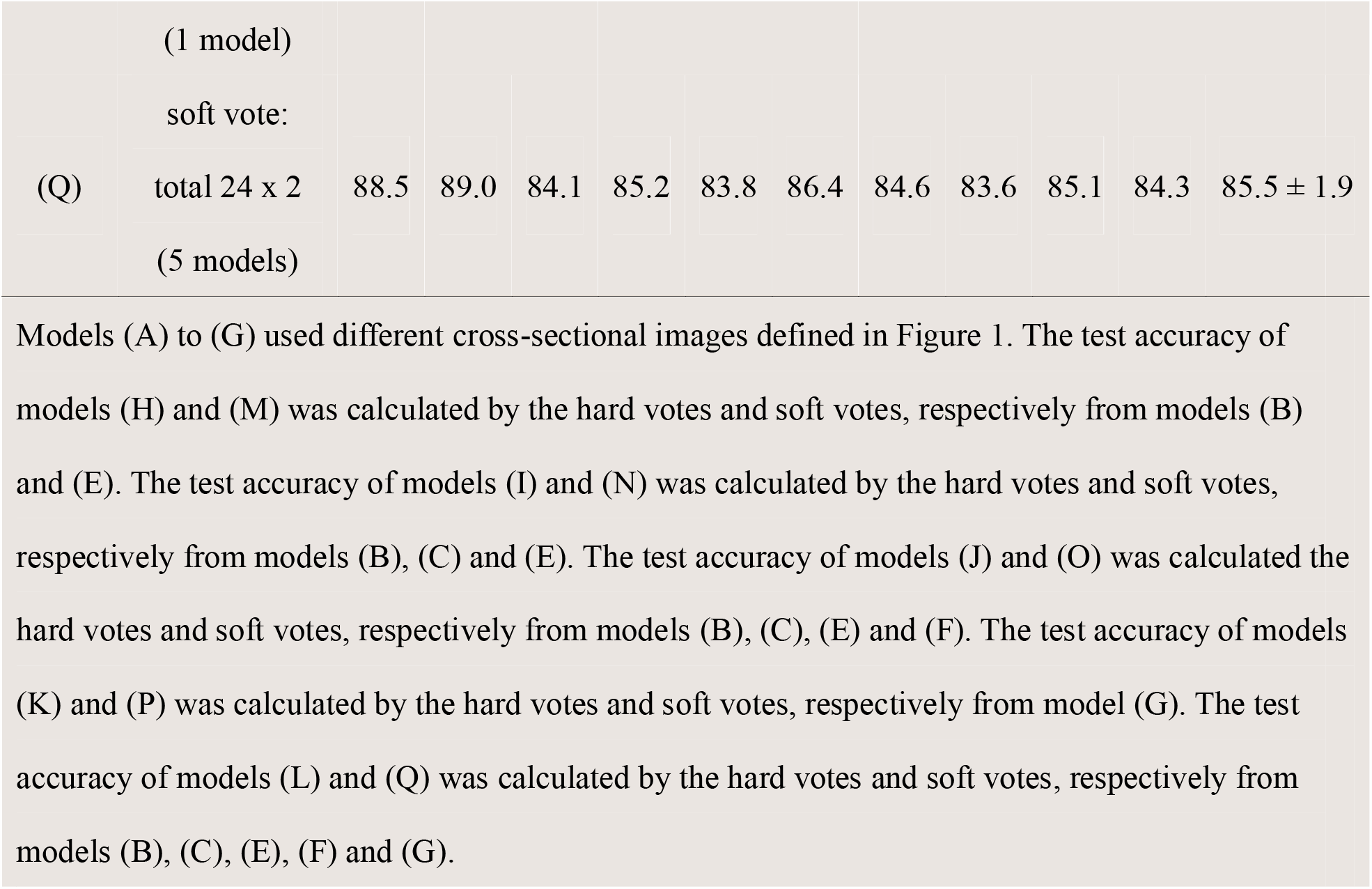
Accuracy for sex prediction using different dataset with 10-fold cross validation.

#### Hard vote & soft vote for deep learning models

Hard vote and soft vote were used to improve the prediction performance by merging the prediction results from different areas at the macula. Table 1 shows the results of a hard vote and soft vote using different model combinations. By using hard vote, the sex prediction accuracy could be up to 85.6 ± 2.1%. Similarly, by using a soft vote, the sex prediction accuracy could be up to 85.5 ± 1.9%.

#### Feature analysis from Grad-CAM and guided Grad-CAM

Next, Grad-CAM overlaid on original OCT images was used to identify the regions that the CNN might have been using to make its predictions. Five representative examples of macular OCT images with their Grad-CAM are shown in Figure 2. To understand more detailed features in the fovea, the Grad-CAM of different layers of ResNet18 were analyzed. In Layer 1, some Grad-CAM focused on the whole retina while some focused on the vitreous, implying that the model’s focus was bounded by the inner retinal surface of the macula. In Layer 2, Grad-CAM focused mainly on the inner limiting membrane and the retinal pigment epithelium layers, which defined both the inner and outer contours of the retina. In Layers 3 and 4, Grad-CAM focused mainly on the fovea. Layer 4 is the last convolutional layer of ResNet18, so the Grad-CAM of Layer 4 should have the highest correlation with the prediction results. According to these results, it is proposed that the deep-learning model used the information of fovea mostly to make the prediction of sex.

**Figure 2.**
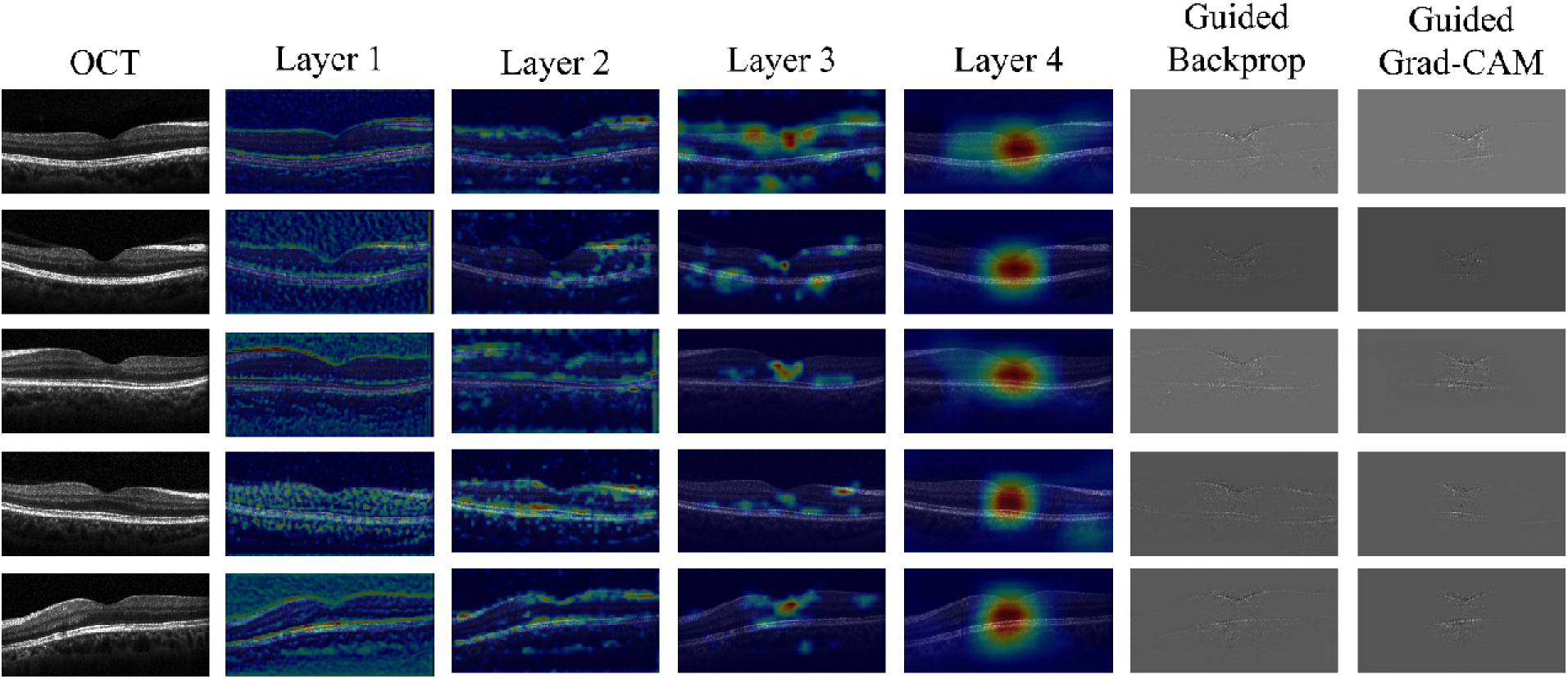
Grad-CAM of different input OCT images. OCT column shows five typical macular OCT images (). These images are predicted correctly by the model. Layer 1, Layer 2, Layer 3, and Layer 4 column show the Grad-CAM in 4 layers of ResNet18, which have a receptive field of 43×43, 99×99, 211×211, 435×435, respectively. Guided Backprop column shows guided backpropagation of OCT. Guided Grad-CAM column is arrived from pointwise multiplying the Grad-CAM by guided backpropagation.

The extracted features by the deep learning models were further analyzed from the guided Grad-CAM. Guided Grad-CAM combines Grad-CAM with existing fine-grained visualization (guided backpropagation) to create a high-resolution class-discriminative visualization. According to the guided Grad-CAM, the inner contour of the fovea was clearly shown, suggesting that the foveal contour is the key point for differentiation between male and female.

Figure 3 shows the Grad-CAM from the 10-fold cross-validation models of one single macular OCT image. Although the 10-fold cross-validation models were trained using different input data as described in Methods, all the 10 Grad-CAMs and guided Grad-CAM had similar presentations as those shown in Figure 2, indicating that the fovea and its contour as the key point for sex prediction is promising. Supplementary 1 shows more Grad-CAM from the 10-fold cross-validation models of one single macular OCT image.

**Figure 3.**
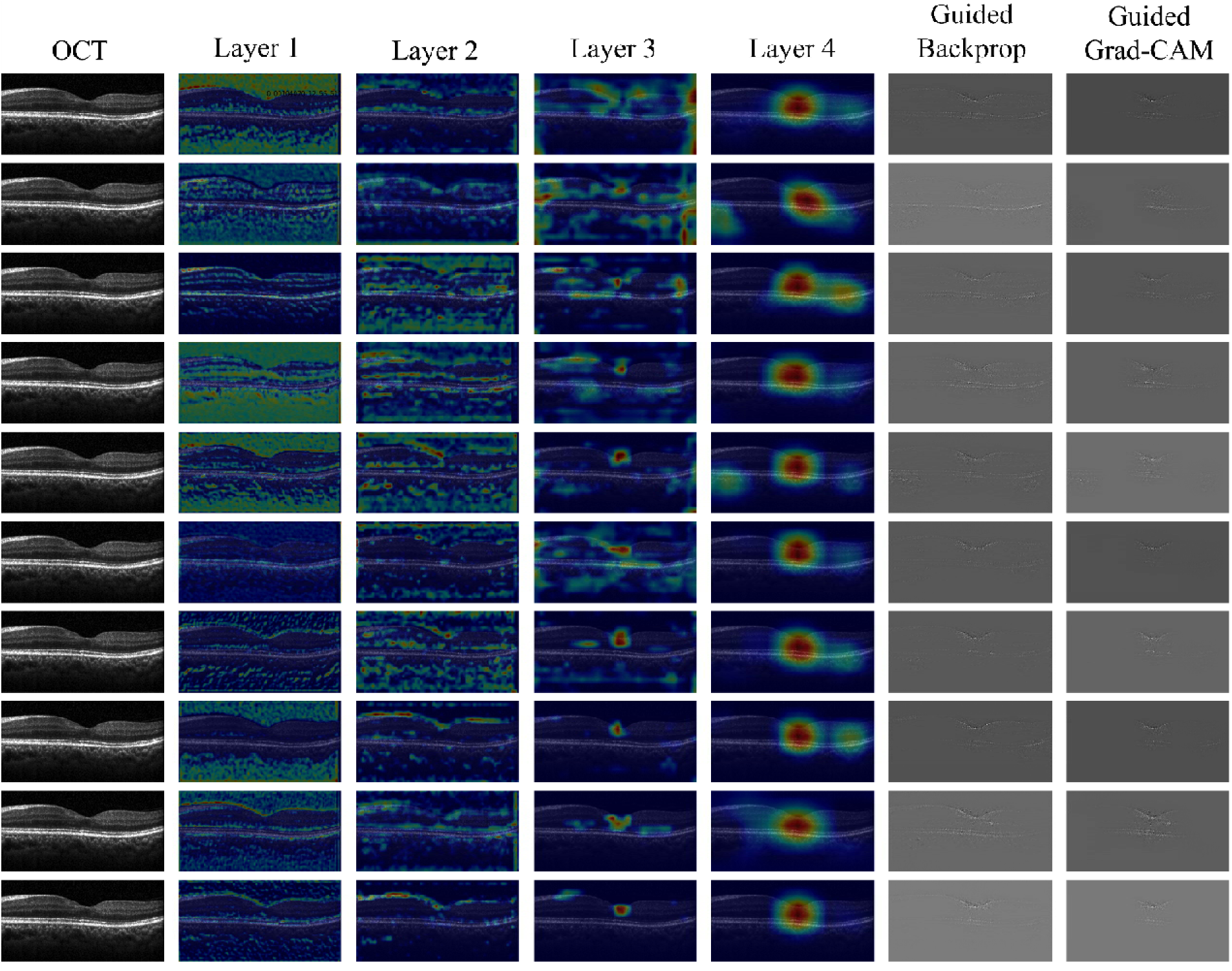
Grad-CAM on one single OCT image with 10-fold cross validation. OCT column shows one macular OCT image (). This image is predicted correctly by all ten cross-validation models. Layer 1, Layer 2, Layer 3, and Layer 4 columns show the Grad-CAM in 4 layers of ResNet18 arrived from ten cross-validation models, which have a receptive field of 43×43, 99×99, 211×211, 435×435, respectively. Guided Backprop column shows guided backpropagation calculated from ten cross-validation models. Guided Grad-CAM column is arrived from pointwise multiplying the Grad-CAM by guided backpropagation.

### Sex prediction using macular thickness

To predict sex by using the macular thickness, 110 persons (56 female, 54 male) with the normal macula in bilateral eyes were collected and the retinal thickness in different areas of the macula were calculated from OCT. A cut-point value of macular thickness in order to differentiate male from female was planned. The best accuracy in differentiating sex was 61.9%, which was obtained using the central 1-mm area of the central fovea (Figure 4a). Furthermore, the data of macular thickness by Melissa et al.^2^ was used to differentiate male from female. The best accuracy was also 61.9%, which was obtained using the temporal perifoveal area (Figure 4b). Both accuracies were poorer than those obtained from deep learning models using macular OCT.

**Figure 4.**
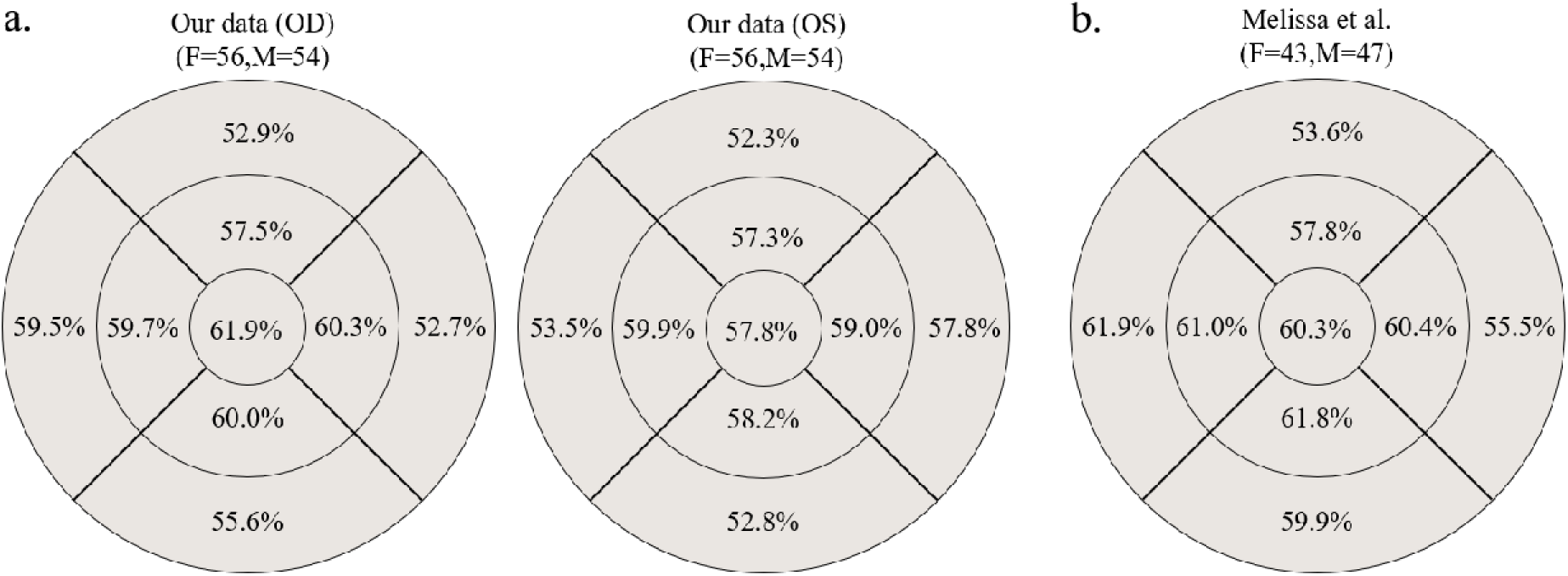
Accuracy for sex prediction using macular thickness. **a**. The accuracy for sex prediction using the macular thickness of 56 female and 54 male using the volume data calculated by the Heidelberg Eye Explorer software. The circles are defined by the ETDRS grid with concentric circles of 1-, 3-, and 6-mm diameters at macula. **b**. The accuracy for sex prediction from the thickness of macula calculated from the data published by Melissa et al.^2^ The details for approach is described in Supplementary 2.

### Sex prediction by deep learning models using infrared fundus photography

The en face image of the macula contains different structural information from the cross-sectional image of the macula. We also used 30° x 30° infrared fundus photography taken during the OCT volume scan to train deep learning models and perform 10-fold cross-validation. The infrared fundus images used were exactly the same individuals as those used for OCT image analysis. Table 2 shows the comparison between infrared fundus photography (384 x 384 pixels, grayscale) and B-scan macular OCT images (256 x 512 pixels) in sex prediction using the images from the same persons. It was found that the accuracy of sex prediction using infrared fundus photography was lower than that using B-scan OCT images; this indicates that B-scan macular OCT images contained more sex-related information than en face infrared fundus photography. Figure 5 is the Grad-CAM models for sex prediction from infrared fundus photography. It shows that the models mainly focused on the optic disc and arcade vessels, which is similar to the results reported by Poplin et al.^8^

**Table 2.** Comparison of sex prediction accuracy between infrared fundus photography and B-scan macular OCT images.

|  |  |  |  |  |  |  |  |  |  |  |  |
| --- | --- | --- | --- | --- | --- | --- | --- | --- | --- | --- | --- |
| Ten-fold validation models |  |  |  |  |  |  |  |  |  |  |  |
| Image | Accuracy (%) |  |  |  |  |  |  |  |  |  |  |
|  | 1 | 2 | 3 | 4 | 5 | 6 | 7 | 8 | 9 | 10 | Mean±SD |
| Infrared fundus | 65.4 | 62.1 | 53.6 | 66.7 | 62.3 | 60.7 | 61.0 | 65.9 | 58.5 | 58.1 | 61.4±4.0 |
| Macular OCT | 72.9 | 75.6 | 73.2 | 74.4 | 71.2 | 74.4 | 73.7 | 70.8 | 71.0 | 73.4 | 73.1±1.6 |

**Figure 5.**
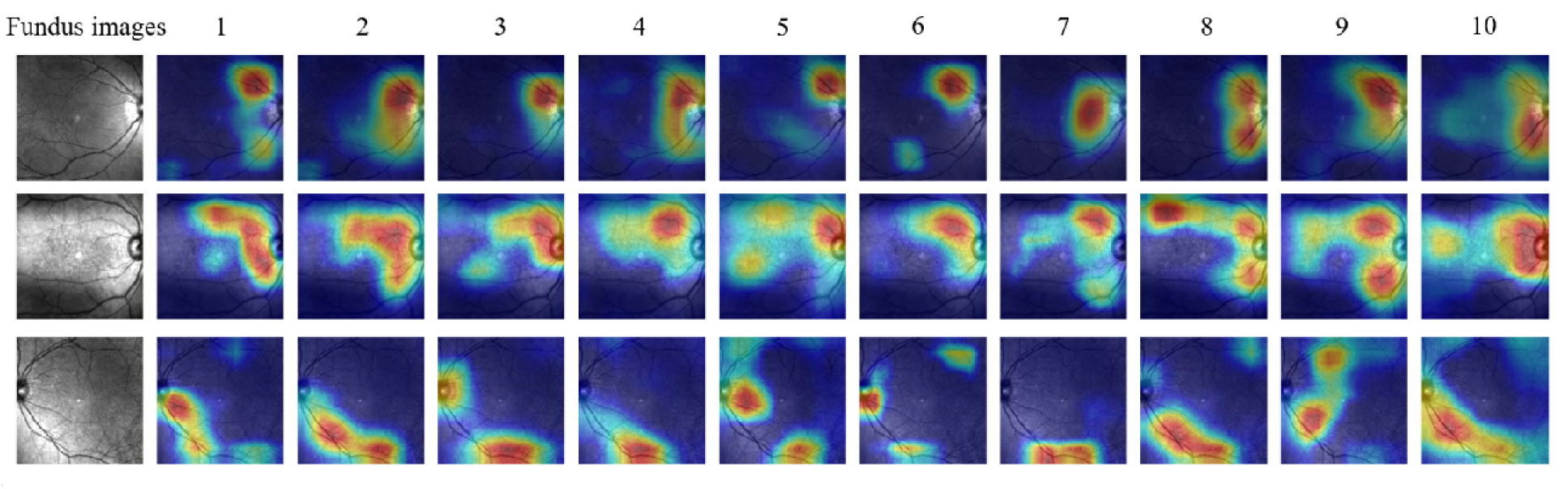
Grad-CAM on 3 infrared fundus images with 10-fold cross validation. The left column shows the infrared fundus photography from three normal eyes. The Grad-CAM of Layer 4 in ResNet18 from 10-fold cross-validation models shows that the hot spots focused mostly on the optic disc and the main arcades of retinal vessels.

### Age prediction using macular OCT images

The deep-learning models were trained to predict the age of patients using the macular OCT images (11^*th*^ and 12^*th*^). The age of patients ranged from 20 to 90 years old. With the 10-fold cross-validation, the average test mean absolute error (MAE) is 5.78 ± 0.29 years (Table 3).

**Table 3.** Mean absolute error for age prediction.

| Table 3 \| Mean absolute error for age prediction |  |  |  |  |  |  |  |  |  |  |
| --- | --- | --- | --- | --- | --- | --- | --- | --- | --- | --- |
| 1 | 2 | 3 | 4 | 5 | 6 | 7 | 8 | 9 | 10 | Mean±SD |
| 6.41 | 5.90 | 6.11 | 5.64 | 5.85 | 5.74 | 5.52 | 5.59 | 5.71 | 5.34 | 5.78±0.29 |
| Mean Absolute Error (MAE) : $\frac{1}{N} \sum \vert Predicted\ age - real\ age\vert $ (unit : year) | | | | | | | | | | |

Figure 6 are images of Grad-CAM from patients of different ages. The images show that the deep learning model predicted age mainly based on the whole layers of the retina, rather than the choroid.

**Figure 6.**
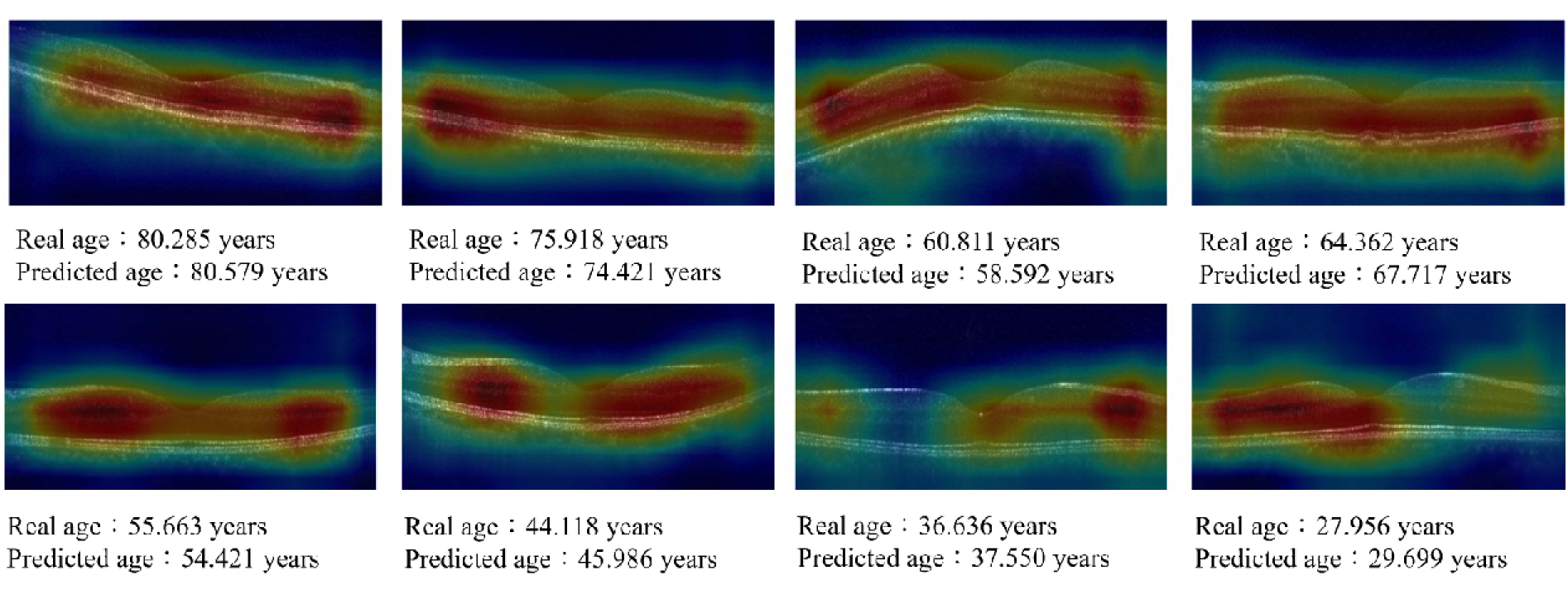
Grad-CAM from the age prediction model. Six examples for the Grad-CAM of OCT images for age prediction. Each image is labeled with the real age and the model’s predicted age.

## Discussion

The present study indicates that macular OCT can be used to predict sex and age via deep learning with good accuracy. For sex prediction, the accuracy could be as high as 85.6%. High accuracy is obtained by using more images in model training as shown in Table 1. Images closer to the central fovea yielded more accurate results, which can be seen from the results of models (A) to (D). A single image that contained the central fovea () out of the 24 macular OCT images for model training and validation provided an accuracy of 73.1% (Table 1 (E)). These results indicate that the central fovea contains more sex-related features than the extrafoveal area. The results from Grad-CAM and guided Grad-CAM also showed that the sex-related difference is focused on the central fovea. Furthermore, differentiation of sex by retinal thickness at different areas of the macula provided an accuracy of only 61.9% at best. These results imply that not only the thickness but also the contour of fovea are different between male and female, which were compatible with the inference from Delori et al.^24^ that the foveal pit is wider in female according to the analysis from fundus reflectometry. Supplementary 3 provides another evidence that the high accuracy rate of the deep learning model for sex prediction was not due to overfitting; the foveal contour is highly sex-related.

The macula is the most important area in the retina because it is in charge of central vision. The sexual difference in macular structure is also an important issue because some macular diseases, such as epiretinal membrane and macular hole, are not equally prevalent between male and female. Using the Grad-CAM and guided Grad-CAM as assistant tools, it was found that the central fovea, especially its contour, is a significant feature of sex. The foveal contour varies largely among normal population; hence little was known about its clinical significance. Poplin et al.^8^ showed that optic nerve head and arcade vessels were highlighted by the Grad-CAM of models for sex prediction using color fundus photography. However, the size of the samples used for model training in their study was much larger (284,335 persons vs 2,844 persons in the present study). The results showed that by using the same number of datasets for training, the accuracy for sex prediction using infrared fundus photography (61.4%) is lower than that using B-scan OCT images (73.1%). This is compatible with the deduction that the foveal contour, which is more clearly shown in B-scan OCT, is much different between male and female. Such results can offer the ophthalmologists an idea to define the foveal contour with a variety of parameters and work on the difference of foveal contour between male and female in order to clarify the sexual correlation with foveal structure-related diseases, such as epiretinal membrane and macular hole. The results of the present study would be helpful for further epidemiological, anatomical, and pathogenic studies of macular structure and its associated diseases.

As for the prediction of age using macular OCT via deep learning models, the MAE for age prediction in this study was 5.78 years for a study population with a mean age of 55.2 ± 15.0 years. Such a result was compatible with the results reported by Poplin et al^8^, in which the MAE for age prediction is 3.26 years for their study population with a mean age of 56.8 ± 8.2 years. This indicates that macular OCT may contain more age-related information since only 6,147 sets of OCT images from 3,134 patients were used for model training and validation in this study, compared to more than 1 million color fundus images from 284,335 patients used in the study by Poplin et al.^8^ Furthermore, the attention maps in their study showed that the model for age prediction focused mostly on retinal vessels. However, the images used in this study for model training and validation are B-scan OCT images, in which only the cross-section image of retinal vessels can be hardly seen. Interestingly, the Grad-CAM in the present study focused on the whole retinal layers of the macula for age prediction. Such a result is compatible with the previous study showing that retinal thickness decreases with age^25^. Although the choroidal thickness also decreases with age^26^, the attention maps in the present study did not focus on choroid. It is possible that the OCT used in this study is spectral-domain OCT without enhanced depth imaging, so the whole choroid could be clearly shown. However, since the variations in retinal thickness and choroidal thickness among the normal population of the same age are high, the thickness itself may not be enough for age differentiation. Further studies are needed for evaluating age-related changes in the retina and choroid.

In conclusion, it was found that deep learning can be used to unveil the sex and age-related difference in macular OCT. Some difference exists in the anatomical structure of the foveal pit between male and female, and this may be related to the sexual difference in the prevalence of some macular structural diseases including epiretinal membrane and macular hole. The present study offers important findings for further studies in the pathogenesis of sex-related or age-related macular structural diseases.

## Methods

### Data acquisition

The data used in this study were obtained from patients who were older than 20 years old and have undergone macular OCT examination for preoperative screening for refractive surgery or cataract surgery. Eyes with vitreous macular traction, epiretinal membrane, macular hole, macular edema, choroidal neovascularization, posterior scleral staphyloma, and any other obvious macular structural abnormalities were excluded from this study.

The size of the B-scan image from the OCT volume scan is 496 pixels ×512 pixels. The axial pixel resolution of the image is 3.9 μm, the lateral pixel resolution is 11 μm, and the distance between each image is 240 μm. This study uses convolutional neural networks to predict age and sex, so the image is labeled with age and sex information obtained from the electronic records.

### Data pre-processing

To make the classification easier, the region of interest was automatically cropped from the original OCT images as shown in Figure 7. First, a certain cropping rectangle was designed in the size of 512 pixels × 256 pixels. Then, from the top of the images, the pixel values within the cropping rectangle are added up and the cropping rectangle moves down one pixel to calculate again until the rectangle reaches the bottom of the image. Finally, the cropped region with the highest total pixel values was chosen for analysis.

**Figure. 7.**
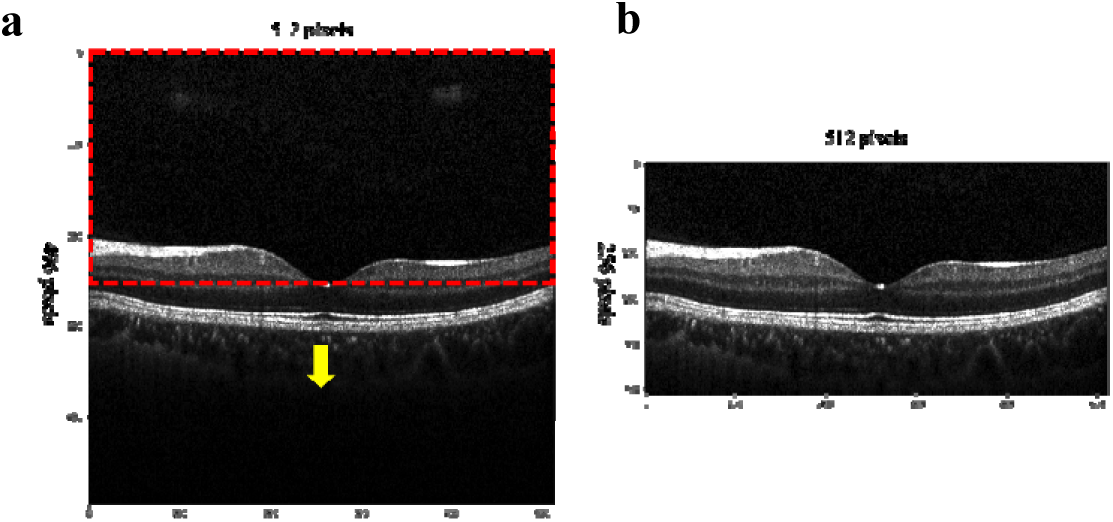
Image cropping method. a. Original OCT image. b. Cropped image after the data pre-processing method.

### Model development

In this study, the pretrained Resnet18 on ImageNet^27^ was used for training. The model’s input was adjusted to be one channel (input size: 512×256), and the output was adjusted to be two ways according to the type of prediction. Supplementary 4 describes the models with more details.

## Code availability

The deep learning models developed in this work employed standard libraries and scripts that are publicly available from https://pytorch.org/.

## Data availability

The authors declare that the main data supporting the results in this study are available, with restrictions, from Yi-Ting Hsieh under request.

## Funding

This study was sponsored by a research grant from the Ministry of Science and Technology of Taiwan (109-2221-E-002-151-).

## Notes

### Competing Interest Statement

The authors have declared no competing interest.

### Author Declarations

This study was approved by the Ethics Committee and Institutional Review Board of National Taiwan University Hospital.

