## Supplementary data for "Prediction of Sex and Age from Macular Optical Coherence Tomography Images and Feature Analysis Using Deep Learning"

**Supplementary 1**

Five examples of grad-CAM from the 10-fold cross validation models of one single macular OCT image.

**Example 1**

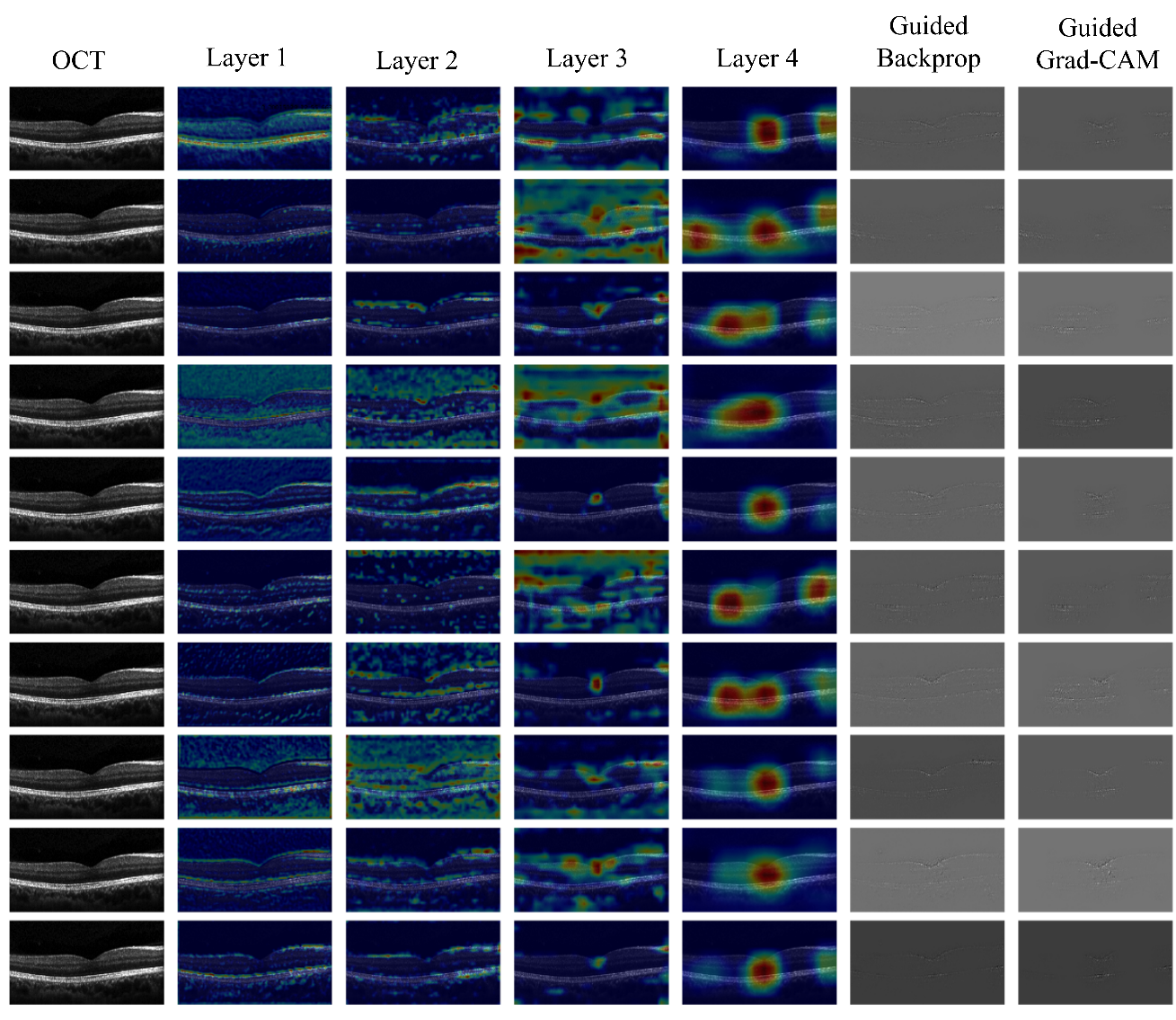

**Example 2**

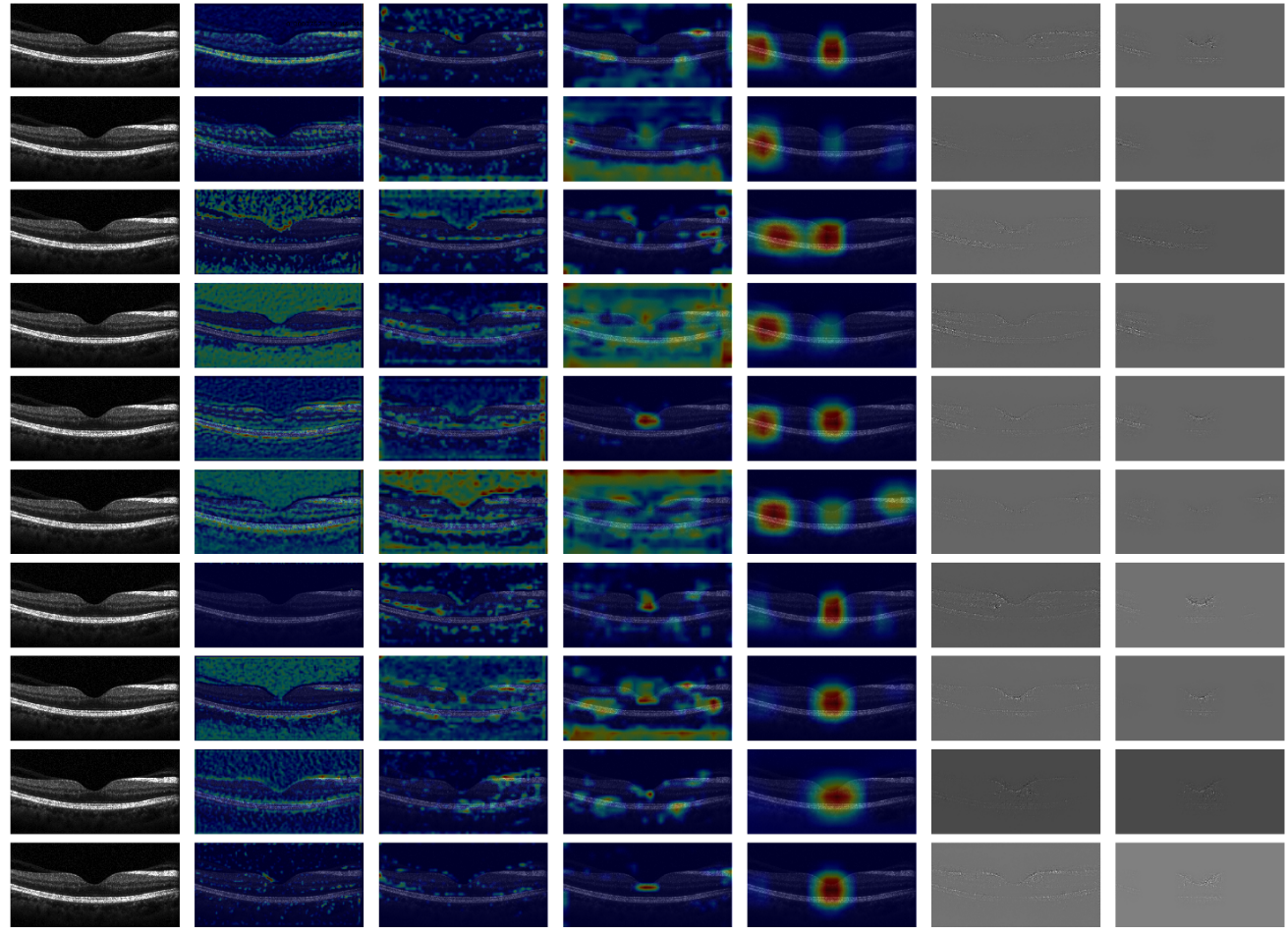

**Example 3**

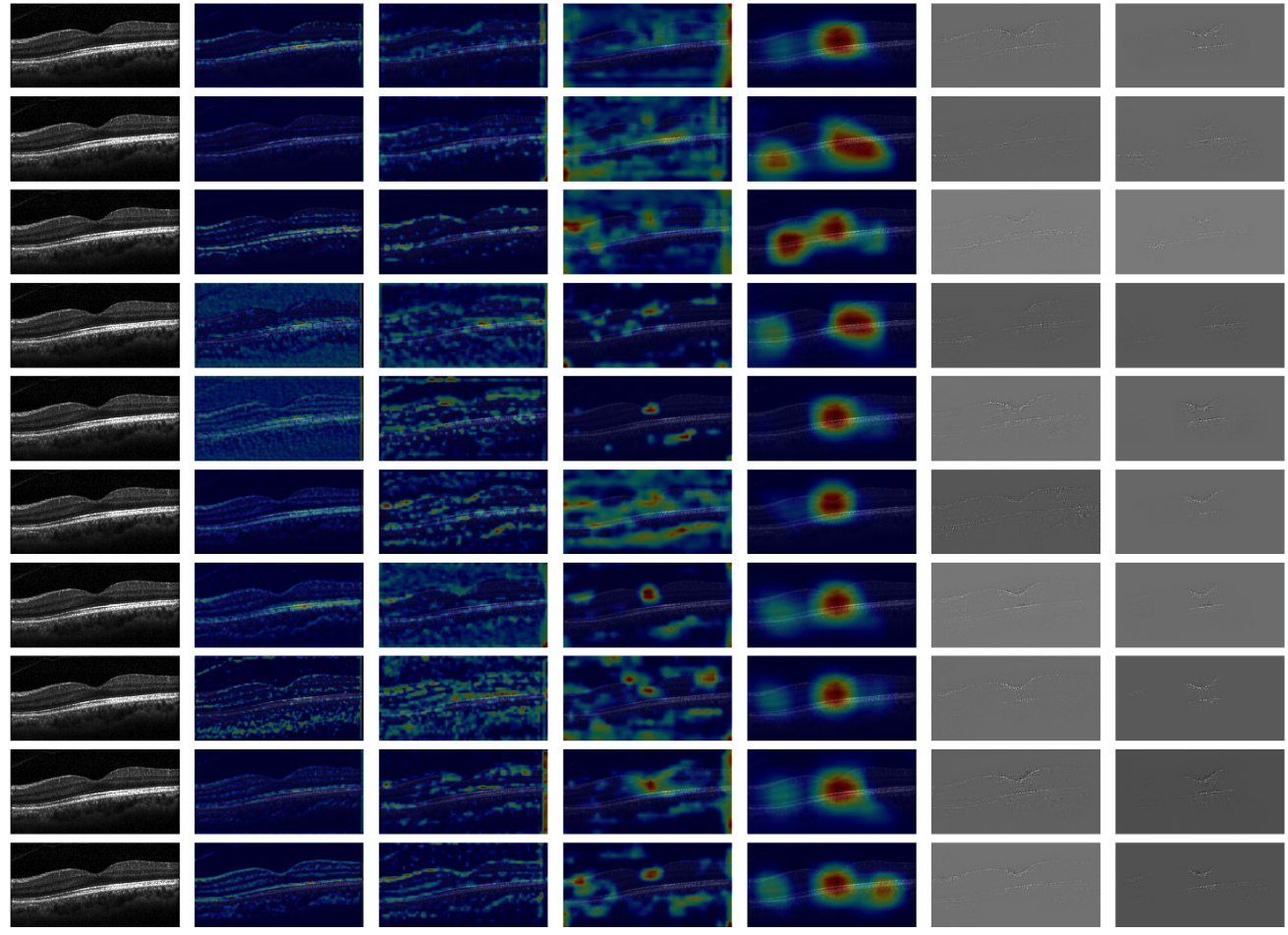

**Example 4**

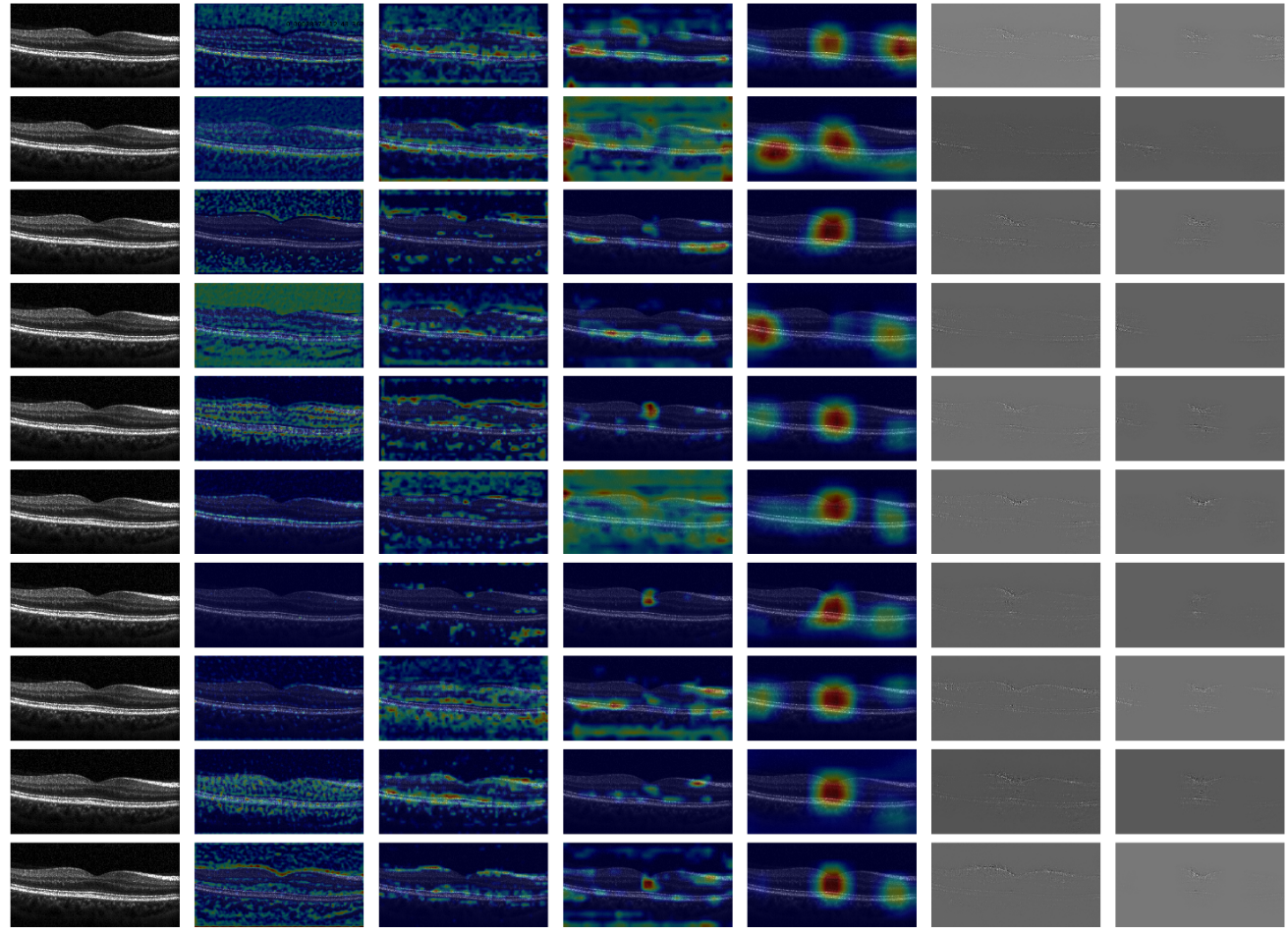

**Example 5**

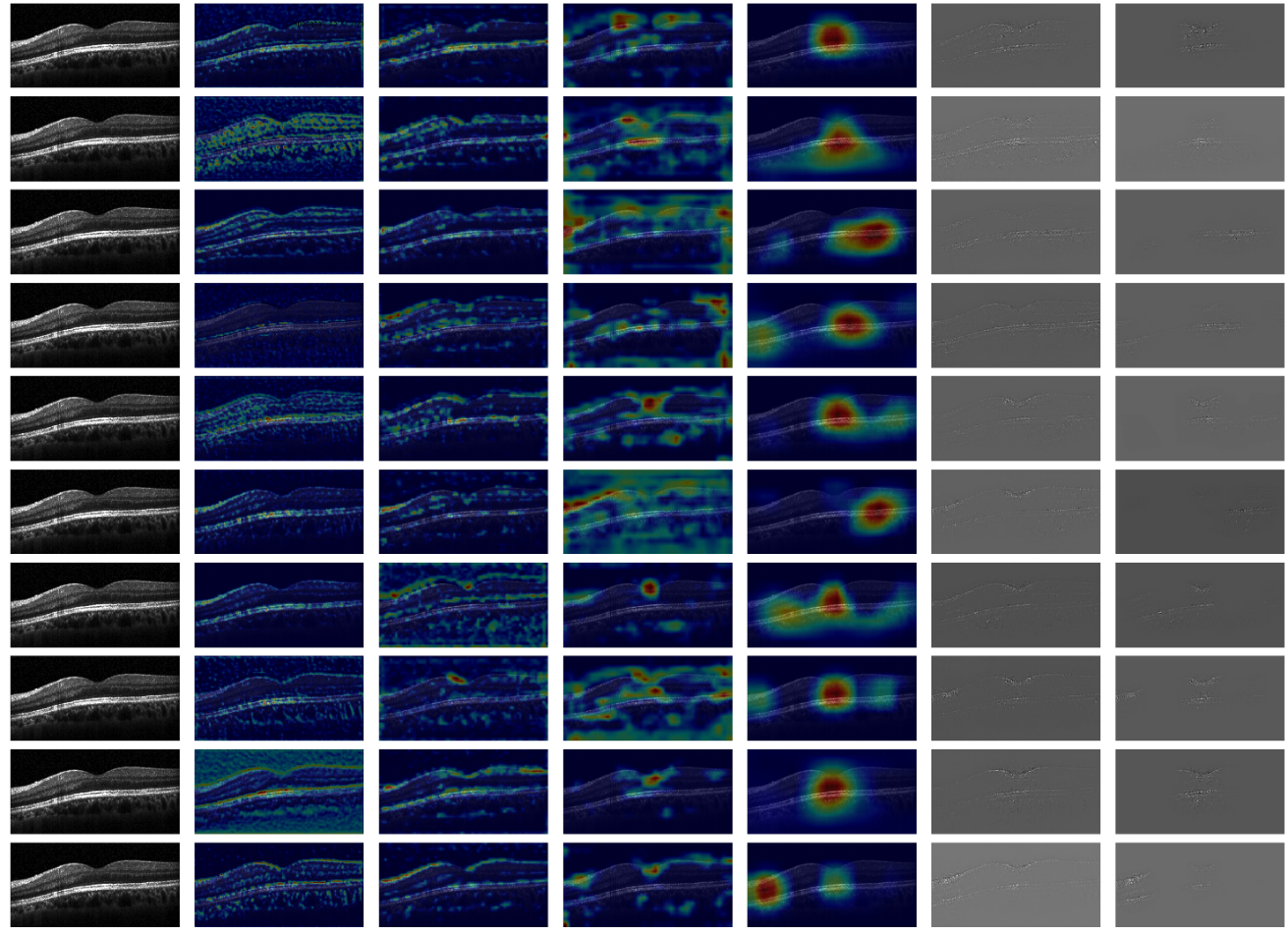

**Supplementary 2**

**Approach for sex prediction by using macular thickness**

The distribution of the macular thickness is fitted with a Gaussian function. For instance, the average central retinal thickness for the female is 253.6 ± 19.3 μm, and the average thickness for male was 264.5 ± 22.8 μm. The red curve and the blue curve in Supplementary Figure 1 were the fitted Gaussian curves for female and male, respectively. Then the distribution curves were used to find a cut point of central retinal thickness that can distinguish sex with the best accuracy.

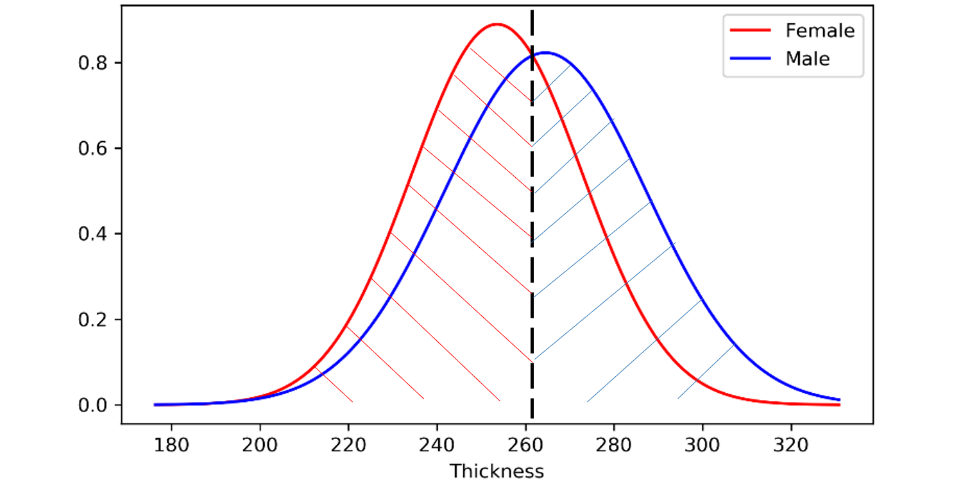

**Supplementary Figure 1 | Thickness distribution of male and female.**

The black imaginary line sets the cut point of central retinal thickness to distinguish male from female. Red slashes show the area separated by the cut point which is the amount of correctly classified female. Blue slashes show the area separated by the boundary which is the amount of correctly classified male.

**Supplementary 3**

To clarify that the high accuracy rate of the deep learning model in differentiating sex was not due to overfitting, we created another parameter to label the sex of the OCT images with a random value (0 or 1) instead of the real sex. Thereafter, the same dataset as used in Table 1(A) was collected to train the new deep learning model for predicting the randomly labeled sex with 10-fold cross validation method. The result showed that the accuracy in such model was only 50.0% (Supplementary Table). This means that the difference between male and female does exist in macular B-scan OCT.

**Table | Prediction accuracy for the real sex and the randomly labeled sex using** **OCT images**

| Predicted parameter | Accuracy (%) | | | | | | | | | | |
| --- | --- | --- | --- | --- | --- | --- | --- | --- | --- | --- | --- |
|  | 1 | 2 | 3 | 4 | 5 | 6 | 7 | 8 | 9 | 10 | Mean±SD |

| Real sex | 76.2 | 78.4 | 74.8 | 79.1 | 73.8 | 77.7 | 77.9 | 71.2 | 74.7 | 75.6 | 75.9±2.3 |
| --- | --- | --- | --- | --- | --- | --- | --- | --- | --- | --- | --- |

| Randomly labeled sex | 45.8 | 50.9 | 47.5 | 48.4 | 49.7 | 53.6 | 50.3 | 48.1 | 52.2 | 53.4 | 50.0±2.6 |
| --- | --- | --- | --- | --- | --- | --- | --- | --- | --- | --- | --- |

**Supplementary 4**

**Details for model development**

For the sex classification, the outputs were two neurons which were the probability of male and female. For the age prediction, the output was one neuron, which was the age. The loss functions for sex classification and age prediction were cross-entropy loss and mean square error loss. The model was trained by the gradient descent method (Adam). In order to avoid the problem of overfitting, L2 regularization, dropout, and data augmentation were adopted:

The L2 regularization technique discourages learning a more complex or flexible model by adding a penalty term. $\frac{1}{n}\sum_{w} w^{2}$ is defined as the Euclidean Norm (or L2 norm) of the weight matrices, which is the average overall squared weight values of a weight matrix. The regularization term is weighted by the scalar $\lambda$ (0.001) divided by two and added to the regular loss function (cross- entropy or mean square error loss). This leads to a new expression for the loss function:

$$L=L_{0}+\frac{\lambda}{2n}\sum_{w} w^{2}$$

$L$ refers to the final loss function, $L_{0}$ refers to the regular loss function, $\lambda$ refers to the scalar constant, $n$ refers to the number of weight, and $w$ refers to the weight value.

A fully connected layer occupies most of the parameters, and hence, neurons develop co-dependency amongst each other during training which curbs the individual power of each neuron leading to over-fitting of training data. Dropout refers to ignoring neurons during the training phase of a certain set of neurons which is chosen at random. In this paper, fully connected layers included dropout with the pulling out rate of 0.5.

Data augmentation is a strategy that significantly increases the diversity of data available for training models. Therefore, during the training process, images were horizontally flipped randomly.

*Grad-CAM*

After training the model, Grad-CAMs**^23^** were used to show the features which were extracted from the model to predict a certain class. The weights of feature maps from the certain layer were the average of class-specific gradient values calculated from the output：

$$w_{k}^{c}=\frac{1}{Z}\sum_{i} \sum_{j} \frac{\partial y^{c}}{\partial A_{ij}^{k}}$$

$w_{k}^{c}$ refers to the weight of $k^{th}$ feature map of class c. $y^{c}$ refers to the output value of class c. $A_{ij}^{k}$ refers to the ($i^{th}$,$j^{th}$) pixel of $k^{th}$ feature map. $Z$ refers to the number of pixels in one feature map.

Grad-CAM is the weighted summation of the feature maps through the activation function of ReLU which deletes the negative value：

$$L_{Grad-CAM}^{c}=ReLU(\sum_{k} w_{k}^{c}A^{k})$$

$L_{Grad-CAM}^{c}$ refers to the Grad-CAM for class c, and $A^{k}$ refers to $k^{th}$ feature map.

Guided Grad-CAM uses the gradients, which are closely related to the importance weights of the corresponding pixels. The importance weight of a pixel evaluates the impact of a change in the pixel on the prediction. The Guided Grad-CAM method, a combination of guided back-propagation and Grad-CAM-is also used to show extracted localized visual features of a single image which has the same resolution as the input image. Since the work for predicting age is not a method of classification, the weights of the feature maps should not be the degree to which the value of the class rises. It is represented as the importance of prediction of that age. Since the higher, the weight, the more important is the feature map. Therefore, the generation of Grad-CAM is to take the absolute value of the calculated weight and then multiply the feature map.

*Hard vote and soft vote*

Several different deep learning models were used for sex prediction in this study. A hard vote is to take the most frequently predicted class of different models for one OCT volume scan as the chosen predicted class. For example, among the prediction results for five different models for one OCT volume scan, three were female and two were male; in this case, the result for the hard vote would be female. A soft vote is to average the probabilities of the predicted classes among different models, and the higher value is taken as the predicted class. For example, the probability for male and female for one OCT volume scan in five different models was (0.12, 0.88), (0.02, 0.98), (0.10, 0.90), (0.46, 0.54), (0.63, 0.37), respectively. The average of the predicted probabilities for male and female was (0.266, 0.734), so in this case, the result of the soft vote would be female.

*Ten-fold cross validation*

To demonstrate that the results are reliable, a 10-fold cross-validation method was executed in all models. The whole data was divided into training, validation, and test data in 10 different ways (Supplementary Figure 2). Every time, one of the data set among the 10 sets was used. The model was trained with training data which contained 80% of the total data. Validation data (10%) then was used to find the best-trained model in the process of training. Finally, test data (10%) was used to examine the model chosen by the validation data.

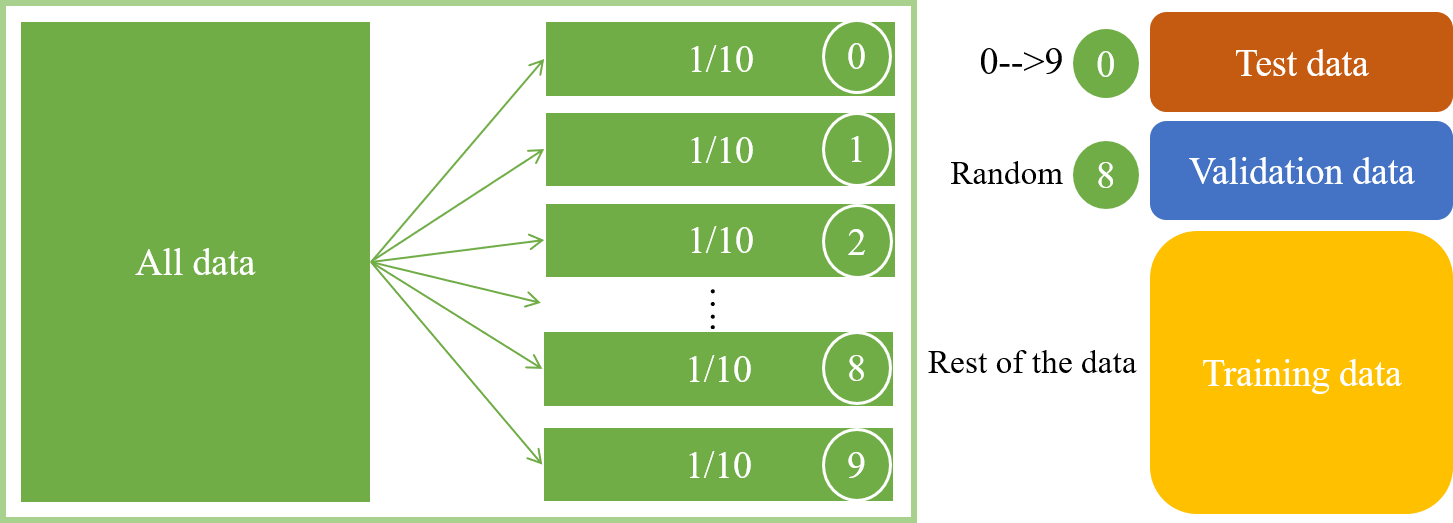

**Supplementary Figure 2 | Ten-fold cross-validation method**

First, the whole data was divided into ten groups (different groups of data contained the same number of images and were all from different people) and assigned each group of data as the test data for ten times. Each time one group of data from the remaining nine groups was randomly selected as the validation data, and the remaining eight groups of data were used as the training data. In this way, the training, validation, and test data used for ten times were all different, and the ratio of the number of images could always be maintained at 8:1:1.
